# Evaluation of GENECUBE^®^ HQ SARS-CoV-2 for anterior nasal samples and saliva samples with a new rapid examination protocol

**DOI:** 10.1101/2021.08.23.21262454

**Authors:** Asami Naito, Yoshihiko Kiyasu, Yusaku Akashi, Akio Sugiyama, Masashi Michibuchi, Yuto Takeuchi, Shigeyuki Notake, Koji Nakamura, Hiroichi Ishikawa, Hiromichi Suzuki

## Abstract

**Introduction:** GENECUBE^®^ is a rapid molecular identification system, and previous studies demonstrated that GENECUBE^®^ HQ SARS-CoV-2 showed excellent analytical performance for the detection of severe acute respiratory syndrome coronavirus-2 (SARS-CoV-2) with nasopharyngeal samples. However, other respiratory samples have not been evaluated.

**Methods:** This prospective comparison between GENECUBE^®^ HQ SARS-CoV-2 and reference real-time reverse transcriptase polymerase chain reaction (RT-PCR) was performed for the detection of SARS-CoV-2 using anterior nasal samples and saliva samples. Additionally, we evaluated a new rapid examination protocol using GENECUBE^®^ HQ SARS-CoV-2 for the detection of SARS-CoV-2 with saliva samples. For the rapid protocol, in the preparation of saliva samples, purification and extraction processes were adjusted, and the total process time was shortened to approximately 35 minutes.

**Results:** For 359 anterior nasal samples, the total-, positive-, and negative concordance of the two assays was 99.7% (358/359), 98.1% (51/52), and 100% (307/307), respectively. For saliva samples, the total-, positive-, and negative concordance of the two assays was 99.6% (239/240), 100% (56/56), and 99.5% (183/184), respectively. With the new protocol, total-, positive-, and negative concordance of the two assays was 98.8% (237/240), 100% (56/56), and 98.4% (181/184), respectively. In all discordance cases, SARS-CoV-2 was detected by additional molecular examinations.

**Conclusion:** GENECUBE^®^ HQ SARS-CoV-2 provided high analytical performance for the detection of SARS-CoV-2 in anterior nasal samples and saliva samples.

## Introduction

For the diagnosis of coronavirus disease 2019 (COVID-19), accurate and rapid laboratory testing is essential. Molecular examination using real-time reverse transcriptase polymerase chain reaction (RT-PCR) has been considered the gold standard for the identification of SARS-CoV-2 [1], and nasopharyngeal samples have been commonly used for the sample examination, which requires high-level personal protective equipment [2]. For COVID-19 testing, anterior nasal samples and saliva samples have been proposed as alternative samples [3], which can be easily obtained from patients.

GENECUBE^®^ (TOYOBO Co., Ltd., Osaka, Japan) is a Qprobe-PCR-based automated rapid molecular identification system that can detect target genes in a short time and simultaneously analyze up to 12 samples and 4 assays in a single examination [4–9]. The system automatically performs molecular examination directly, including preparation of the reaction mixtures, and amplification and detection of target genes, in 30 minutes.

GENECUBE^®^ HQ SARS-CoV-2 (TOYOBO Co., Ltd.) is the GENECUBE^®^ reagent for detecting the SARS-CoV-2 gene in clinical samples. This reagent was approved in October, 2021. We previously evaluated the performance of the assay using 1065 nasopharyngeal samples [10]. Compared with the reference RT-PCR assay, the overall positive- and negative concordance rates were 100.0% (95% confidence interval [CI]: 93.4%–100.0%) and 99.7% (95% CI: 99.1%–99.9%), respectively. All discordant samples were GENECUBE^®^ HQ SARS-CoV-2-positive and reference RT-PCR-negative, and SARS-CoV-2 was detected by another molecular assay [10]. During the previous evaluation, samples other than nasopharyngeal samples were not used.

In the present study, we evaluated the diagnostic performance of the GENECUBE^®^ HQ SARS-CoV-2 using anterior nasal samples and saliva samples. Additionally, we evaluated a new rapid examination protocol using GENECUBE^®^ HQ SARS-CoV-2 for the detection of SARS-CoV-2.

## Materials and Methods

The current study was performed at a drive-through PCR center in Tsukuba Medical Center Hospital (TMCH) in Tsukuba, Ibaraki Prefecture, Japan, which intensively performed sample collecting and PCR analysis with nasopharyngeal samples in the Tsukuba district [10]. Anterior nasal samples were prospectively collected from COVID-19-suspected or COVID-19-confirmed patients between 11 May 2021 and 5 July 2021. Saliva samples were prospectively collected from COVID-19-confirmed patients between 21 April 2021 and 13 May 2021. All of anterior nasal samples and saliva samples from COVID-19-confirmed patients were obtained on the same day of nasopharyngeal sample collection.

Anterior nasal samples and saliva samples were simultaneously examined using GENECUBE^®^ HQ SARS-CoV-2 (GENECUBE examination) and reference RT-PCR, and the concordance of the two methods was evaluated.

The ethics committee of Tsukuba Medical Center Hospital approved the present study (approval number: 2020-066) for both anterior nasal sampling and positive saliva samples, informed consent was obtained from patients for their participation in the respective part of the current research. This study was performed in line with the principles of the Declaration of Helsinki and adheres to the STARD reporting guidelines.

For negative saliva samples, residual frozen saliva samples collected during SARS-CoV-2 active screening at hospitalization in Tsukuba Medical Center Hospital were used for the current research after anonymization.

### Sample Collection

For anterior nasal samples, a nasopharyngeal-type flocked swab (Copan Italia SpA, Brescia, Italy) was inserted to a 2-cm depth in one nasal cavity, rotated five times, and held in place for 5 seconds. The swab samples were then diluted in 3 mL of UTM^™^ (Copan Italia SpA) immediately after sampling, and the UTM^™^ was then transferred to a microbiology laboratory located next to the drive-through sampling facility of the PCR center.

After arrival, purification, and ribonucleic acid (RNA) extraction were performed with magLEAD (Precision System Science Co., Ltd., Chiba, Japan) with 200-µL of fresh anterior nasal samples. RNA was eluted in 100-µL, which was used for the GENECUBE^®^ examination and the reference RT-PCR examination. All saliva samples were stored at −80°C and were purified with magLEAD after preparation (Fig. 1). All of the GENECUBE^®^ examinations and reference RT-PCR examinations were performed simultaneously on the same day.

**Fig. 1.**
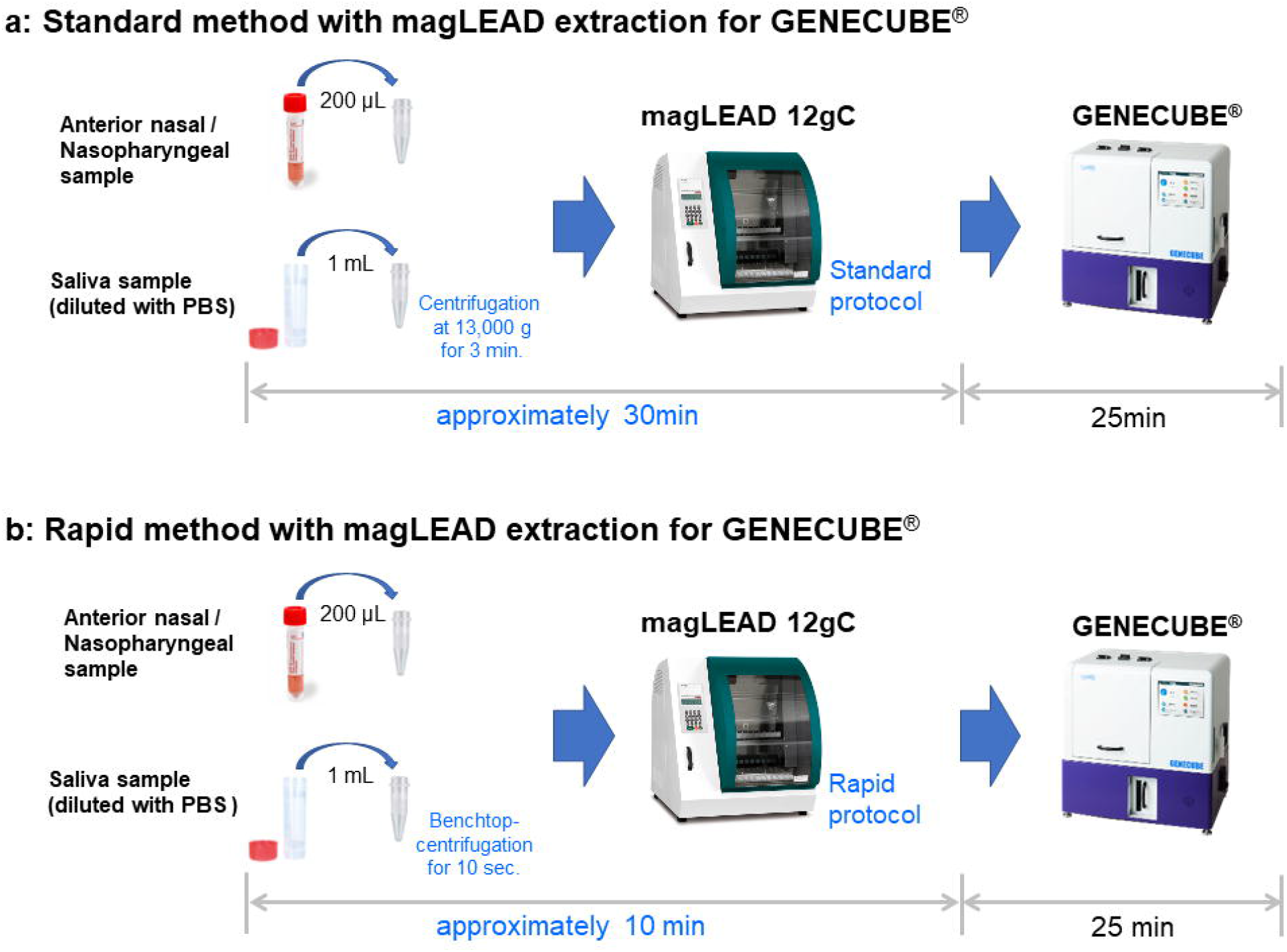
Workflow of the two extraction methods for the GENECUBE^®^ assay in this study. The rapid method with magLEAD extraction (b) newly developed in this study takes as little as 10 min for viral RNA extraction, while the standard method (a) takes approximately 30 min. For the rapid protocol, in the preparation of saliva samples, purification and extraction processes were adjusted, and the total process time was shortened. *PBS* Phosphate-buffered saline.

### GENECUBE^®^ Examination with magLEAD and Discrepancy Analysis

The sample used for the GENECUBE^®^ examination analysis of SARS-CoV-2 was also used for the RT-PCR assays. All assays were performed with the previously described magLEAD conditions [10] (Fig. 1). In addition to the standard protocol with magLEAD purification, a rapid protocol created by Hiromichi Suzuki, TOYOBO Co., Ltd. and Precision System Science Co., Ltd. were evaluated for saliva samples and samples for limit of detection (LOD) analysis. For the rapid protocol, in the preparation of saliva samples, purification and extraction processes were adjusted, and the total process time was shortened to approximately 10 minutes.

If discordance was recognized between GENECUBE^®^ and the reference RT-PCR, an additional evaluation was performed with Xpert^®^ Xpress SARS-CoV-2 and GeneXpert^®^ (Cepheid Inc., Sunnyvale, CA, USA) [12] analyses for anterior nasal samples, and with an RT-PCR with LightMix^®^ Modular SARS-CoV (COVID19) E-gene (Roche Diagnostics KK) [13] for saliva samples along with re-evaluation with the NIID RT-PCR method.

### Reference Real-time RT-PCR Method

Reference RT-PCR examinations were performed with purified samples using a method developed by the National Institute of Infectious Diseases (NIID), Japan, for SARS-CoV-2 [11]. Briefly, 5 μL of the extracted RNA was used for one-step quantitative RT-PCR with the THUNDERBIRD^®^ Probe One-step qRT-PCR kit (TOYOBO Co., Ltd) and the LightCycler^®^ 96 Real-time PCR System (Roche Diagnostics KK, Basel, Switzerland). A duplicate analysis for N2 genes was performed for the evaluation of SARS-CoV-2.

### Evaluation of the Limit of Detection (LOD) for GENECUBE^®^ HQ SARS-CoV-2 with Nasopharyngeal Samples and Saliva Samples

To evaluate the LOD for GENECUBE^®^ HQ SARS-CoV-2, we made four different concentrations of samples (2500 copies/mL, 1000 copies/mL, 500 copies/mL, 250 copies/mL) with SARS-CoV-2 reference material (AccuPlex^™^ SARS-CoV-2 Reference Material Kit, SeraCare; SeraCare Life Sciences, Inc., Milford, MA, USA) and matrix (UTM^™^; three pooled nasopharyngeal samples and two pooled saliva samples). In total, six samples were made at each concentration. The GENECUBE^®^ examination was performed four times, and the reference RT-PCR was performed twice for each sample.

## Statistical Analyses

The positive-, negative-, and total concordance rates of the GENECUBE^®^ examinations compared with the reference RT-PCR were calculated using the Clopper and Pearson methods with 95% confidence intervals. All calculations were conducted using the R 3.3.1 software program (The R Foundation, Vienna, Austria).

## Results

### Evaluation of LOD for the Reference RT-PCR and GENECUBE^®^ with SARS-CoV-2 Reference Material and Pooled Negative Samples

The details of the results of the LOD evaluation for the three SARS-CoV-2 detection methods with SARS-CoV-2 reference material and pooled negative samples are listed in Table 1 and summarized in Table 2.

**Table 1.**
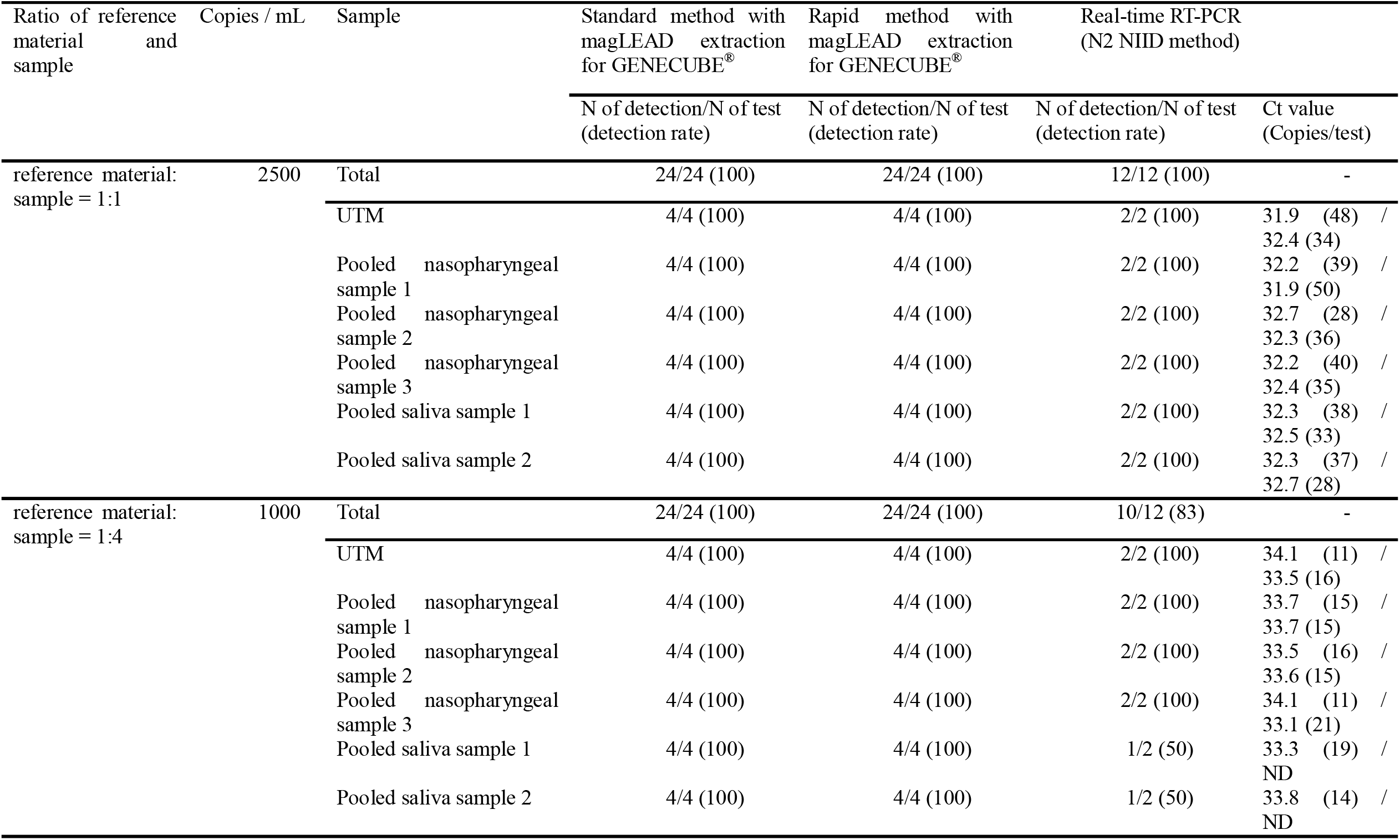
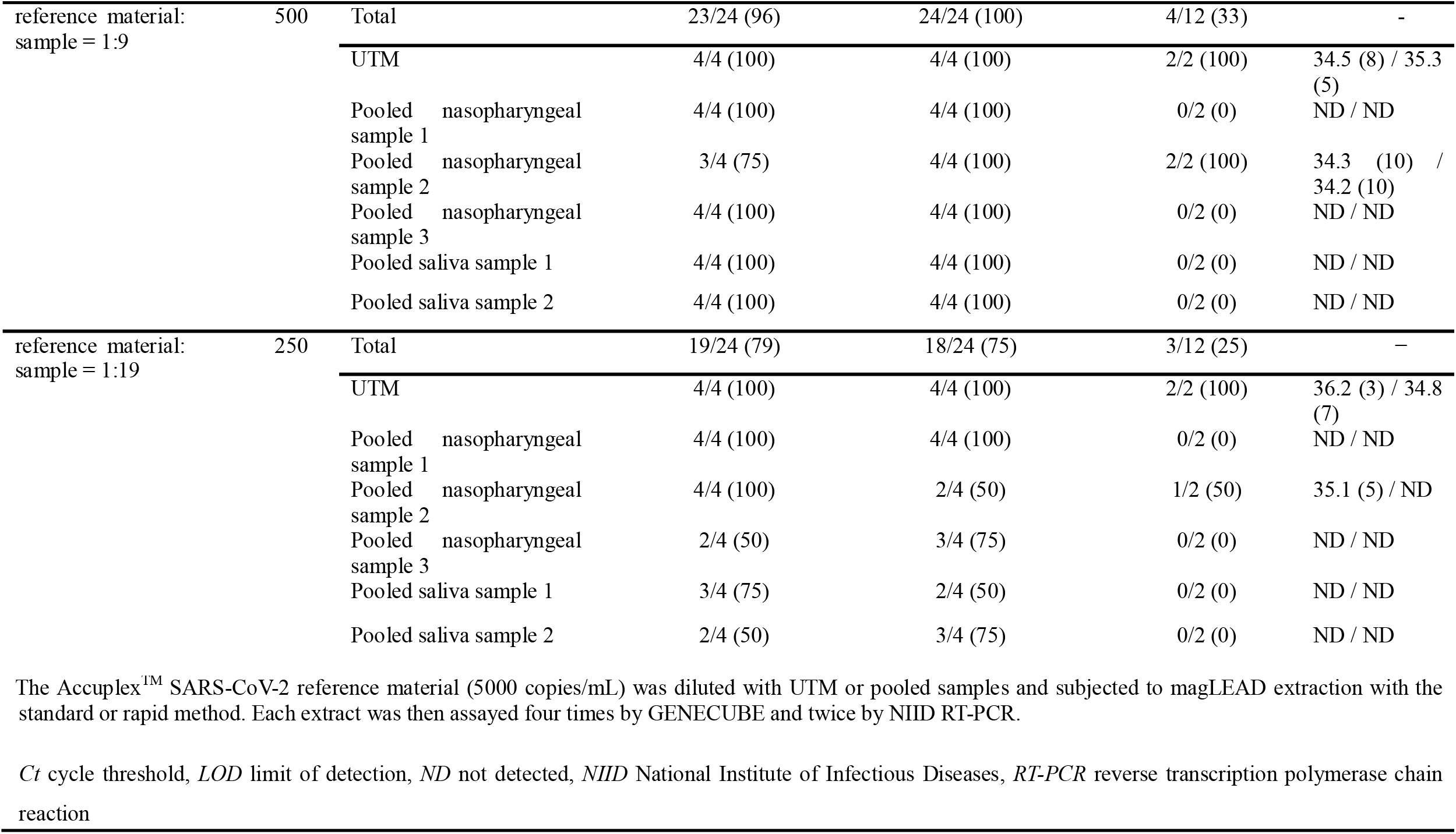
Detailed results of the LOD test for three SARS-CoV-2 detection methods

| Ratio of reference material and sample | Copies / mL | Sample | Standard method with magLEAD extraction for GENECUBE® | Rapid method with magLEAD extraction for GENECUBE® | Real-time RT-PCR (N2 NIID method) |  |
| --- | --- | --- | --- | --- | --- | --- |
|  |  |  | N of detection/N of test (detection rate) | N of detection/N of test (detection rate) | N of detection/N of test (detection rate) | Ct value (Copies/test) |
| reference material: sample = 1:1 | 2500 | Total | 24/24 (100) | 24/24 (100) | 12/12 (100) | - |
|  |  | UTM | 4/4 (100) | 4/4 (100) | 2/2 (100) | 31.9 (48) / 32.4 (34) |
|  |  | Pooled nasopharyngeal sample 1 | 4/4 (100) | 4/4 (100) | 2/2 (100) | 32.2 (39) / 31.9 (50) |
|  |  | Pooled nasopharyngeal sample 2 | 4/4 (100) | 4/4 (100) | 2/2 (100) | 32.7 (28) / 32.3 (36) |
|  |  | Pooled nasopharyngeal sample 3 | 4/4 (100) | 4/4 (100) | 2/2 (100) | 32.2 (40) / 32.4 (35) |
|  |  | Pooled saliva sample 1 | 4/4 (100) | 4/4 (100) | 2/2 (100) | 32.3 (38) / 32.5 (33) |
|  |  | Pooled saliva sample 2 | 4/4 (100) | 4/4 (100) | 2/2 (100) | 32.3 (37) / 32.7 (28) |
| reference material: sample = 1:4 | 1000 | Total | 24/24 (100) | 24/24 (100) | 10/12 (83) | - |
|  |  | UTM | 4/4 (100) | 4/4 (100) | 2/2 (100) | 34.1 (11) / 33.5 (16) |
|  |  | Pooled nasopharyngeal sample 1 | 4/4 (100) | 4/4 (100) | 2/2 (100) | 33.7 (15) / 33.7 (15) |
|  |  | Pooled nasopharyngeal sample 2 | 4/4 (100) | 4/4 (100) | 2/2 (100) | 33.5 (16) / 33.6 (15) |
|  |  | Pooled nasopharyngeal sample 3 | 4/4 (100) | 4/4 (100) | 2/2 (100) | 34.1 (11) / 33.1 (21) |
|  |  | Pooled saliva sample 1 | 4/4 (100) | 4/4 (100) | 1/2 (50) | 33.3 (19) / ND |
|  |  | Pooled saliva sample 2 | 4/4 (100) | 4/4 (100) | 1/2 (50) | 33.8 (14) / ND |
| reference material:<br>sample = 1:9 | 500 | Total | 23/24 (96) | 24/24 (100) | 4/12 (33) | - |
|  |  | UTM | 4/4 (100) | 4/4 (100) | 2/2 (100) | 34.5 (8) / 35.3 (5) |
|  |  | Pooled nasopharyngeal sample 1 | 4/4 (100) | 4/4 (100) | 0/2 (0) | ND / ND |
|  |  | Pooled nasopharyngeal sample 2 | 3/4 (75) | 4/4 (100) | 2/2 (100) | 34.3 (10) / 34.2 (10) |
|  |  | Pooled nasopharyngeal sample 3 | 4/4 (100) | 4/4 (100) | 0/2 (0) | ND / ND |
|  |  | Pooled saliva sample 1 | 4/4 (100) | 4/4 (100) | 0/2 (0) | ND / ND |
|  |  | Pooled saliva sample 2 | 4/4 (100) | 4/4 (100) | 0/2 (0) | ND / ND |
| reference material:<br>sample = 1:19 | 250 | Total | 19/24 (79) | 18/24 (75) | 3/12 (25) | - |
|  |  | UTM | 4/4 (100) | 4/4 (100) | 2/2 (100) | 36.2 (3) / 34.8 (7) |
|  |  | Pooled nasopharyngeal sample 1 | 4/4 (100) | 4/4 (100) | 0/2 (0) | ND / ND |
|  |  | Pooled nasopharyngeal sample 2 | 4/4 (100) | 2/4 (50) | 1/2 (50) | 35.1 (5) / ND |
|  |  | Pooled nasopharyngeal sample 3 | 2/4 (50) | 3/4 (75) | 0/2 (0) | ND / ND |
|  |  | Pooled saliva sample 1 | 3/4 (75) | 2/4 (50) | 0/2 (0) | ND / ND |
|  |  | Pooled saliva sample 2 | 2/4 (50) | 3/4 (75) | 0/2 (0) | ND / ND |

The Accuplex™ SARS-CoV-2 reference material (5000 copies/mL) was diluted with UTM or pooled samples and subjected to magLEAD extraction with the standard or rapid method. Each extract was then assayed four times by GENEcube and twice by NIID RT-PCR.
*Ct* cycle threshold, *LOD* limit of detection, *ND* not detected, *NIID* National Institute of Infectious Diseases, *RT-PCR* reverse transcription polymerase chain reaction

**Table 2.** Summary of the LOD test results for three SARS-CoV-2 detection methods

| Sample<br>(Copies / mL) | Standard method with<br>magLEAD extraction for<br>GENECUBE® | Rapid method with magLEAD<br>extraction for GENECUBE® | Real-time RT-PCR<br>(N2 NIID method) |
| --- | --- | --- | --- |
|  | N of detection/N of test<br>(detection rate) | N of detection/N of test<br>(detection rate) | N of detection/N of test<br>(detection rate) |
| 2500 | 24/24 (100) | 24/24 (100) | 12/12 (100) |
| 1000 | 24/24 (100) | 24/24 (100) | 10/12 (83) |
| 500 | 23/24 (96) | 24/24 (100) | 4/12 (33) |
| 250 | 19/24 (79) | 18/24 (75) | 3/12 (25) |

*LOD* limit of detection, *N* number, *NIID* National Institute of Infectious Diseases, *RT-PCR* reverse transcription polymerase chain reaction

The reference NIID real-time RT-PCR method showed positive results for all UTM-based samples (range: 250–5000 copies/mL), while the detection rate was 100% down to 1000 copies/mL for pooled nasopharyngeal samples and down to 2500 copies/mL for pooled saliva-based samples. None of the 500 copies/mL of pooled saliva-based samples and 250 copies/mL of pooled saliva-based samples were detected by the NIID real-time RT-PCR method.

The standard protocols with magLEAD and GENECUBE^®^ showed positive results for all UTM-based samples. The detection rate was 100% down to 1000 copies/mL for pooled nasopharyngeal-based samples and down to 500 copies/mL for pooled saliva-based samples. The detection rate of 500 copies/mL pooled nasopharyngeal-based samples was 91.7% (11/12).

For the rapid protocol with magLEAD and GENECUBE^®^, the method showed positive results for all UTM-based samples. The detection rate was 100% down to 500 copies/mL for pooled nasopharyngeal-based samples and pooled saliva-based samples.

#### Comparison of the Reference RT-PCR and GENECUBE^®^ for the Detection of SARS-CoV-2 with Anterior Nasal Samples

In this study, we prospectively evaluated 359 fresh anterior nasal samples, including 59 samples with positive SARS-CoV-2 results for simultaneously collected nasopharyngeal samples (cycle threshold (Ct) < 20, n = 40; Ct ≥ 20 to < 30, n =16; Ct ≥ 30, n =3) (Supplementary Table 1). Of the 359 anterior nasal samples, 298 (83.0%) were obtained from asymptomatic patients.

The comparison of the reference RT-PCR and GENECUBE^®^ (standard protocol) for the detection of SARS-CoV-2 with anterior nasal samples is summarized in Table 3. The total-, positive-, and negative concordance of the two assays was 99.7% (358/359), 98.1% (51/52), and 100% (307/307), respectively. The viral load of the discordant sample (sample #42; supplementary table 1) was 1 copy/test by the reference RT-PCR and it was positive by an Xpert^®^ Xpress SARS-CoV-2 and GeneXpert^®^.

**Table 3.** Concordance rate of the GENECUBE^®^ HQ SARS-CoV-2 with real-time RT-PCR for anterior nasal samples

|  |  | Real-time RT-PCR (N2 NIID method) |  |
| --- | --- | --- | --- |
|  |  | Positive | Negative |
| Standard method with magLEAD<br>extraction for GENECUBE® | Positive | 51 | 0 |
|  | Negative | 1* | 307 |
| Positive concordance rate (%) |  | 98.1 (89.7–100) |  |
| Negative concordance rate (%) |  | 100 (98.8–100) |  |
| Total concordance rate (%) |  | 99.7 (98.5–100) |  |

*NIID* National Institute of Infectious Diseases, *RT-PCR* reverse transcription polymerase chain reaction
Data in parentheses are 95% confidence intervals.
\*The discordant sample was tested by Xpert® Xpress SARS-CoV-2 and GeneXpert® and was positive.

#### Comparison of the Reference RT-PCR and GENECUBE^®^ for the Detection of SARS-CoV-2 with Saliva Samples

For the comparison between the reference RT-PCR and GENECUBE^®^ for the detection of SARS-CoV-2 with 240 frozen saliva samples, the evaluation of the standard protocol with magLEAD and GENECUBE^®^ is described in Table 4-a, and the evaluation of the rapid protocol with magLEAD and GENECUBE^®^ is described in Table 4-b. The result of one sample was invalid initially by both GENECUBE^®^ examinations, and the sample required four-fold dilution with lysis buffer for the GENECUBE^®^ examinations.

**Table 4-a.** Concordance rate of the standard method with magLEAD extraction for GENECUBE^®^ with real-time RT-PCR for saliva samples

|  |  | Real-time RT-PCR (N2 NIID method) |  |
| --- | --- | --- | --- |
|  |  | Positive | Negative |
| Standard method with magLEAD<br>extraction for GENECUBE® | Positive | 56 | 1* |
|  | Negative | 0 | 183 |
| Positive concordance rate (%) |  | 100 (93.6–100) |  |
| Negative concordance rate (%) |  | 99.5 (97.0–100) |  |
| Total concordance rate (%) |  | 99.6 (97.7–100) |  |

*NIID* National Institute of Infectious Diseases, *RT-PCR* reverse transcription polymerase chain reaction
Data in parentheses are 95% confidence intervals.
\*The discordant sample was tested by real-time RT-PCR with Roche LightMix Modular SARS and Wuhan CoV E-gene and was positive.

**Table 4-b.** Concordance rate of the rapid method with magLEAD extraction for GENECUBE^®^ with real-time RT-PCR for saliva samples

|  |  | Real-time RT-PCR (N2 NIID method) |  |
| --- | --- | --- | --- |
|  |  | Positive | Negative |
| Rapid method with magLEAD extraction for GENECUBE<sup>®</sup> | Positive | 56 | 3* |
|  | Negative | 0 | 181 |
| Positive concordance rate (%) |  | 100 (93.6–100) |  |
| Negative concordance rate (%) |  | 98.4 (95.3–99.7) |  |
| Total concordance rate (%) |  | 98.8 (96.4–99.7) |  |

NIID National Institute of Infectious Diseases, *RT-PCR* reverse transcription polymerase chain reaction
Data in parentheses are 95% confidence intervals.
\*The discordant samples were tested by real-time RT-PCR with Roche LightMix Modular SARS and Wuhan CoV E-gene and all were positive.

For the evaluation of the standard protocol with magLEAD and GENECUBE^®^ (Table 4-a), the total-, positive-, and negative concordance of the two assays was 99.6% (239/240), 100% (56/56), and 99.5% (183/184), respectively. During the analysis with the standard protocol with magLEAD and GENECUBE^®^, re-test was required for two samples owing to an invalid result with the first GENECUBE^®^ analysis. Regarding the single sample with a positive GENECUBE^®^ result and negative reference RT-PCR result (Table 5), a positive result was obtained using another RT-PCR with LightMix^®^ Modular SARS-CoV (COVID19) E-gene (n = 2, Ct = 33.6 and 33.7) and by reference RT-PCR with residual purified samples made using the rapid protocol (n = 1, Ct = 33.5). For the evaluation of the rapid protocol with magLEAD and GENECUBE^®^ (Table 4-b), the total-, positive-, and negative concordance of the two assays was 98.8% (237/240), 100% (56/56), and 98.4% (181/184), respectively. Regarding the three samples with a positive GENECUBE^®^ result and negative reference RT-PCR result (Table 5), a positive result was obtained in one of the three samples as described for the evaluation of the standard protocol. However, the other two samples were negative with the additional two RT-PCR methods. For these two samples, positive results were obtained by RT-PCR with LightMix^®^ Modular SARS-CoV (COVID19) E-gene for purified samples made by the rapid protocol (n = 8, 4/8 for one sample, 1/8 for another sample). In the evaluation of E-gene analysis with purified samples with QIAamp^®^ Viral RNA Mini Kit (Qiagen, Valencia, CA, USA), RNA extraction by the manual method was negative for all assays (n = 8; 0/8 for both samples).

**Table 5.** Detailed data for the three cases with discordant findings between the three SARS-CoV-2 detection methods for saliva samples

| Sample No. | GENECUBE® |  | Real-time RT-PCR (N2 NIID method) | Real-time RT-PCR (Roche LightMix Modular SARS and Wuhan CoV E-gene) |  |  |
| --- | --- | --- | --- | --- | --- | --- |
|  | Standard method with magLEAD | Rapid method with magLEAD | Standard method with magLEAD | Standard method with magLEAD | Rapid method with magLEAD | QIAamp® Viral RNA Mini Kit |
| #11 | – | + | 0/2 (0) | 0/2 (0) | 4/8 (50) | 0/8 (0) |
| #31 | + | + | 0/2 (0) | 2/2 (100) | NA | NA |
| #39 | – | + | 0/2 (0) | 0/2 (0) | 1/8 (12.5) | 0/8 (0) |

+ positive, – negative, *NIID* National Institute of Infectious Diseases, *RT-PCR*, reverse transcription polymerase chain reaction, *NA*, Not applicable Values indicate N of detection/N of test (detection rate).

## Discussion

During the analytical evaluation with 359 anterior nasal samples and 240 saliva samples, the GENECUBE^®^ evaluation with GENECUBE^®^ HQ SARS-CoV-2 showed high concordance rates compared with the reference RT-PCR. Compared with the standard protocol, the rapid protocol showed better sensitivity for the detection of SARS-CoV-2 in saliva samples. LOD evaluation indicated that the GENECUBE^®^ examination with the standard protocol and the rapid protocol can detect lower viral load samples than reference RT-PCR.

Saliva has been considered a good alternative for the detection of SARS-CoV-2 in COVID-19 patients [3] and has been widely used in COVID-19 practice. Among rapid molecular identification systems, GeneXpert^®^ showed good analytical performance for the detection of SARS-CoV-2 with saliva samples [14]; however, the application of saliva to rapid molecular identification systems remains a challenge owing to saliva’s viscosity and RT-PCR inhibition. In our current study, we used 60 saliva samples with positive nasopharyngeal sample results for SARS-CoV-2 (Supplementary Table 2), and the rapid protocol detected SARS-CoV-2 in most samples (98.3%; 59/60). The rapid protocol can detect SARS-CoV-2 with high performance in approximately 35 minutes with saliva samples, and the protocol is expected to have clinical utility.

Anterior nasal samples are also a good alternative method for COVID-19 sampling [3]. The method has been reported as less painful and induced fewer coughs or sneezes compared with nasopharyngeal sampling [15]. The application of self-collected anterior nasal sampling has also been reported [16]. In the current study, the analytical performance of the GENECUBE^®^ examination was almost identical to the reference RT-PCR. However, there were seven negative results using anterior nasal samples, which were obtained from patients with positive nasopharyngeal samples (Supplementary Table 1). The detection rate was 88.1% (52/59), which was inferior to saliva samples. In this study, we obtained anterior nasal samples from one nasal cavity; however, it might be necessary to obtain samples from both nasal cavities [17].

There are several limitations in this study that should be mentioned. First, the current research was performed at a PCR center in Japan. The influence of LODs of the GENECUBE^®^ evaluation for genetic variants of SARS-CoV-2 was not evaluated in this study. Second, the current evaluated rapid protocol showed excellent performance for the detection of SARS-CoV-2; however, the sample size was insufficient to conclude that the protocol can be used in clinical practice; additional evaluation in studies with large samples is required. Third, the current GENECUBE^®^ examination can analyze only 12 samples at a single run, and the amplification curve is not displayed. The system must be improved for better examination.

In conclusion, the GENECUBE^®^ examination with GENECUBE^®^ HQ SARS-CoV-2 provided high analytical performance for the detection of SARS-CoV-2 in anterior nasal samples and saliva samples.

## Data Availability

Some restrictions will apply.

## Acknowledgments

We thank Mrs. Yoko Ueda, Mrs. Mio Matsumoto, Mrs. Asami Nakayama, and the staff in the Department of the Clinical Laboratory of Tsukuba Medical Center Hospital for their intensive support of this study. We thank all of the medical institutions for providing their patients’ clinical information.

## Author Contributions

AN, AS, HS drafted the manuscript. YA performed the statistical analyses. HS supervised the project. AN, AS, MM contributed to the execution of molecular assays. YK, YA, YT, SN, KN, and HI supported the preparation of this manuscript. All authors contributed to the writing of the final manuscript.

## Supplemental Files

**Supplementary Table 1** Results of SARS-Cov-2 detection for anterior nasal samples

**Supplementary Table 2** Results of SARS-Cov-2 detection for saliva samples

