## Supplementary Table 1 and 2 for "Evaluation of GENECUBE^®^ HQ SARS-CoV-2 for anterior nasal samples and saliva samples with a new rapid examination protocol"

*ｍedRxiv*

Asami Naito^1^ • Yoshihiko Kiyasu^2,3^ • Yusaku Akashi^2,4^ • Akio Sugiyama^5^ • Masashi Michibuchi^5^ • Yuto Takeuchi^2,3^ • Shigeyuki Notake^6^ • Koji Nakamura^6^ • Hiroichi Ishikawa^7^ • Hiromichi Suzuki^2,3,8^

^1^Tsukuba i-Laboratory LLP, 2-1-17, Amakubo, Tsukuba, Ibaraki 305-0005, Japan

^2^Division of Infectious Diseases, Department of Medicine, Tsukuba Medical Center Hospital, 1-3-1 Amakubo, Tsukuba, Ibaraki 305-8558, Japan

^3^Department of Infectious Diseases, University of Tsukuba Hospital, 2-1-1 Amakubo, Tsukuba, Ibaraki 305-8576, Japan

^4^Akashi Internal Medicine Clinic, 3-1-63 Asahigaoka, Kashiwara, Osaka 582-0026, Japan

^5^Diagnostic System Department, TOYOBO Co., Ltd., 2-2-8, Dojima Hama, Kita-ku, Osaka, 530-8230, Japan

^6^Department of Clinical Laboratory, Tsukuba Medical Center Hospital, 1-3-1 Amakubo, Tsukuba, Ibaraki 305-8558, Japan

^7^Department of Respiratory Medicine, Tsukuba Medical Center Hospital, 1-3-1 Amakubo, Tsukuba, Ibaraki 305-8558, Japan

^8^Department of Infectious Diseases, Faculty of Medicine, University of Tsukuba, 1-1-1 Tennodai, Tsukuba, Ibaraki 305-8575, Japan

* Correspondence to:

Hiromichi Suzuki, MD, PhD

**Supplementary Table 1** Results of SARS-Cov-2 detection for anterior nasal samples

| Anterior nasal samples | | | | | Nasopharyngeal samples collected from the same patient simultaneously | | |
| --- | --- | --- | --- | --- | --- | --- | --- |
| Sample No. | GENECUBE_®_  (Standard method with magLEAD) | Real-time RT-PCR  (N2 NIID method) | | | GENECUBE_®_  (Standard method with magLEAD) | Real-time RT-PCR  (N2 NIID method) | |
|  |  | Ct | Ct | Copies/test |  | Ct | Copies/test |
| #1 | + | 18.1 | 18.1 | 171,250 | + | 13.7 | 3,828,000 |
| #2 | + | 20.3 | 20.3 | 99,765 | + | 11.3 | 18,800,000 |
| #3 | + | 22.8 | 22.8 | 18,340 | + | 19.1 | 145,600 |
| #4 | + | 29.8 | 29.7 | 162 | + | 16.5 | 745,000 |
| #5 | + | 37.7 | ND | 1 | + | 24.5 | 5,063 |
| #6 | + | 22.3 | 22.3 | 24,840 | + | 14.1 | 7,360,000 |
| #7 | + | 24.1 | 24.1 | 7,601 | + | 17.8 | 588,500 |
| #8 | + | 23.3 | 23.3 | 9,821 | + | 13.2 | 4,672,000 |
| #9 | + | 23.1 | 23.2 | 10,725 | + | 16.5 | 601,000 |
| #10 | + | 19.9 | 19.9 | 202,700 | + | 20.0 | 103,400 |
| #11 | + | 24.6 | 24.6 | 6,764 | + | 25.0 | 3,959 |
| #12 | + | 18.6 | 18.6 | 753,200 | + | 16.4 | 3,162,000 |
| #13 | + | 14.8 | 14.8 | 6,096,500 | + | 15.4 | 7,636,000 |
| #14 | + | 16.6 | 16.5 | 1,804,500 | + | 17.4 | 1,569,000 |
| #15 | + | 33.1 | 32.6 | 31 | + | 31.1 | 68 |
| #16 | + | 17.8 | 17.8 | 707,300 | + | 13.3 | 11,340,000 |
| #17 | + | 26.4 | 26.4 | 1,810 | + | 19.9 | 135,000 |
| #18 | + | 18.1 | 18.1 | 574,000 | + | 16.5 | 1,258,000 |
| #19 | + | 20.9 | 20.9 | 41,260 | + | 13.0 | 5,015,000 |
| #20 | + | 34.2 | 34.3 | 10 | + | 29.1 | 283 |
| #21 | + | 31.1 | 31.1 | 76 | + | 25.7 | 2,654 |
| #22 | + | 18.4 | 18.4 | 268,950 | + | 19.3 | 151,000 |
| #23 | + | 19.9 | 19.9 | 102,100 | + | 20.5 | 72,690 |
| #24 | + | 16.3 | 16.3 | 3,603,000 | + | 14.0 | 19,650,000 |
| #25 | + | 18.5 | 18.5 | 303,200 | + | 15.8 | 1,734,000 |
| #26 | + | 19.5 | 19.5 | 154,100 | + | 17.8 | 472,300 |
| #27 | + | 27.1 | 27.2 | 818 | + | 19.1 | 106,000 |
| #28 | + | 34.2 | 33.8 | 14 | + | 24.8 | 3,465 |
| #29 | + | 29.7 | 29.8 | 172 | + | 24.3 | 7,297 |
| #30 | + | 21.6 | 21.5 | 49,900 | + | 19.3 | 236,400 |
| #31 | + | ND | 37.7 | 1 | + | 22.8 | 14,010 |
| #32 | + | 16.7 | 16.8 | 605,850 | + | 14.7 | 2,212,000 |
| #33 | + | 25.9 | 26.0 | 2,380 | + | 23.4 | 13,770 |
| #34 | + | 16.8 | 16.8 | 1,257,500 | + | 15.5 | 2,982,000 |
| #35 | + | 23.1 | 23.2 | 12,155 | + | 15.6 | 1,466,000 |
| #36 | + | 18.8 | 18.8 | 189,600 | + | 15.2 | 1,939,000 |
| #37 | + | 24.2 | 24.2 | 6,399 | + | 17.6 | 408,900 |
| #38 | + | 26.7 | 26.8 | 1,356 | + | 17.4 | 863,300 |
| #39 | + | 25.5 | 25.5 | 3,146 | + | 17.3 | 641,800 |
| #40 | + | 25.5 | 25.5 | 3,239 | + | 14.1 | 5,296,000 |
| #41 | + | 20.7 | 20.8 | 70,325 | + | 16.5 | 1,102,000 |
| #42 | - | ND | 36.8 | 1 | + | 31.2 | 76 |
| #43 | + | 25.3 | 25.3 | 4,674 | + | 16.6 | 1,999,000 |
| #44 | + | 27.1 | 27.2 | 1,274 | + | 19.7 | 224,400 |
| #45 | + | 19.0 | 19.0 | 383,000 | + | 12.4 | 38,160,000 |
| #46 | + | 29.2 | 29.4 | 301 | + | 22.8 | 25,310 |
| #47 | + | 18.9 | 18.9 | 366,500 | + | 18.3 | 537,900 |
| #48 | + | 25.4 | 25.3 | 1,869 | + | 16.3 | 233,500 |
| #49 | + | 27.8 | 27.9 | 507 | + | 18.9 | 57,760 |
| #50 | + | 30.5 | 30.6 | 119 | + | 23.4 | 5,236 |
| #51 | + | 22.2 | 22.2 | 31,420 | + | 15.0 | 4,452,000 |
| #52 | + | 33.4 | 32.8 | 19 | + | 20.8 | 81,770 |
|  | - | ND | ND | ND | + | 20.0 | 83,610 |
|  | - | ND | ND | ND | + | 19.7 | 171,900 |
|  | - | ND | ND | ND | + | 31.7 | 54 |
|  | - | ND | ND | ND | + | 16.1 | 1,402,000 |
|  | - | ND | ND | ND | + | 19.8 | 200,500 |
|  | - | ND | ND | ND | + | 29.4 | 220 |
|  | - | ND | ND | ND | + | 26.1 | 1,281 |

*Ct* cycle threshold, *ND* not detected, *NIID* National Institute of Infectious Diseases, *RT-PCR* reverse transcription polymerase chain reaction

**Evaluation of GENECUBE HQ SARS-CoV-2 for anterior nasal samples and saliva samples with a new rapid examination protocol**

*ｍedRxiv*

Asami Naito^1^ • Yoshihiko Kiyasu^2,3^ • Yusaku Akashi^2,4^ • Akio Sugiyama^5^ • Masashi Michibuchi^5^ • Yuto Takeuchi^2,3^ • Shigeyuki Notake^6^ • Koji Nakamura^6^ • Hiroichi Ishikawa^7^ • Hiromichi Suzuki^2,3,8^

^1^Tsukuba i-Laboratory LLP, 2-1-17, Amakubo, Tsukuba, Ibaraki 305-0005, Japan

^2^Division of Infectious Diseases, Department of Medicine, Tsukuba Medical Center Hospital, 1-3-1 Amakubo, Tsukuba, Ibaraki 305-8558, Japan

^3^Department of Infectious Diseases, University of Tsukuba Hospital, 2-1-1 Amakubo, Tsukuba, Ibaraki 305-8576, Japan

^4^Akashi Internal Medicine Clinic, 3-1-63 Asahigaoka, Kashiwara, Osaka 582-0026, Japan

^5^Diagnostic System Department, TOYOBO Co., Ltd., 2-2-8, Dojima Hama, Kita-ku, Osaka, 530-8230, Japan

^6^Department of Clinical Laboratory, Tsukuba Medical Center Hospital, 1-3-1 Amakubo, Tsukuba, Ibaraki 305-8558, Japan

^7^Department of Respiratory Medicine, Tsukuba Medical Center Hospital, 1-3-1 Amakubo, Tsukuba, Ibaraki 305-8558, Japan

^8^Department of Infectious Diseases, Faculty of Medicine, University of Tsukuba, 1-1-1 Tennodai, Tsukuba, Ibaraki 305-8575, Japan

* Correspondence to:

Hiromichi Suzuki, MD, PhD

**Supplementary Table 2** Results of SARS-Cov-2 detection for saliva samples

| Saliva samples | | | | | | Nasopharyngeal samples collected from the same patient simultaneously | | |
| --- | --- | --- | --- | --- | --- | --- | --- | --- |
| Sample No. | GENECUBE_®_  (Standard method with magLEAD) | GENECUBE_®_  (Rapid method with magLEAD) | Real-time RT-PCR  (N2 NIID method) | | | GENECUBE_®_  (Standard method with magLEAD) | Real-time RT-PCR  (N2 NIID method) | |
|  |  |  | Ct | Ct | Copies/test |  | Ct | Copies/test |
| #1 | + | + | 18.7 | 19.0 | 113,330 | + | 12.3 | 56,410,000 |
| #2 | + | + | 14.5 | 14.5 | 1,311,050 | + | 14.2 | 4,314,000 |
| #3 | + | + | 32.5 | 32.3 | 51 | + | 23.7 | 6,357 |
| #4 | + | + | 21.8 | 22.0 | 19,160 | + | 14.8 | 2,534,000 |
| #5 | + | + | 23.2 | 23.3 | 9,013 | + | 20.2 | 87,800 |
| #6 | + | + | 30.8 | 30.9 | 121 | + | 19.2 | 123,600 |
| #7 | + | + | 35.2 | ND | 8 | + | 29.4 | 238 |
| #8 | + | + | 15.6 | 15.7 | 694,100 | + | 10.7 | 14,010,000 |
| #9 | + | + | 22.4 | 21.9 | 16,485 | + | 18.3 | 217,800 |
| #10 | + | + | 31.9 | 31.9 | 68 | + | 22.9 | 11,010 |
| #11 | - | + | ND | ND | ND | + | 20.1 | 71,130 |
| #12 | + | + | 14.3 | 14.5 | 1,417,500 | + | 14.9 | 960,000 |
| #13 | + | + | 15.8 | 15.9 | 623,700 | + | 14.7 | 3,024,000 |
| #14 | + | + | 31.1 | ND | 87 | + | 18.6 | 253,200 |
| #15 | + | + | 22.7 | 22.2 | 13,780 | + | 15.5 | 5,599,000 |
| #16 | + | + | 25.4 | 25.0 | 2,952 | + | 15.1 | 2,382,000 |
| #17 | - | - | ND | ND | ND | + | 29.7 | 172 |
| #18 | + | + | 28.9 | 28.9 | 359 | + | 23.6 | 6,551 |
| #19 | + | + | 24.3 | 24.6 | 4,578 | + | 27.9 | 548 |
| #20 | + | + | 14.6 | 14.6 | 1,257,850 | + | 16.8 | 546,900 |
| #21 | + | + | 28.3 | 28.4 | 498 | + | 30.4 | 120 |
| #22 | + | + | 19.7 | 20.0 | 63,540 | + | 15.3 | 1,477,000 |
| #23 | + | + | 32.7 | 33.1 | 38 | + | 20.7 | 32,500 |
| #24 | + | + | 27.2 | 27.3 | 915 | + | 21.2 | 46,920 |
| #25 | + | + | 16.9 | 16.9 | 334,250 | + | 11.2 | 6,972,000 |
| #26 | + | + | 28.5 | 29.0 | 401 | + | 17.1 | 646,000 |
| #27 | + | + | 30.4 | 30.3 | 162 | + | 17.1 | 366,800 |
| #28 | + | + | 29.1 | 28.9 | 340 | + | 21.2 | 35,070 |
| #29 | + | + | 24.7 | 24.9 | 3,771 | + | 15.8 | 991,000 |
| #30 | + | + | 20.0 | 20.2 | 54,105 | + | 14.1 | 15,390,000 |
| #31 | + | + | ND | ND | ND | + | 23.2 | 7,789 |
| #32 | + | + | 26.1 | 26.3 | 1,672 | + | 19.4 | 152,000 |
| #33 | + | + | 21.0 | 20.5 | 37,410 | + | 17.1 | 671,500 |
| #34 | + | + | 15.0 | 15.1 | 959,300 | + | 18.1 | 148,800 |
| #35 | + | + | 20.8 | 21.0 | 34,915 | + | 16.5 | 379,100 |
| #36 | + | + | 19.9 | 19.3 | 69,525 | + | 15.6 | 1,236,000 |
| #37 | + | + | 23.0 | 23.2 | 9,915 | + | 19.7 | 248,600 |
| #38 | + | + | 29.0 | 28.6 | 388 | + | 25.7 | 1,782 |
| #39 | - | + | ND | ND | ND | + | 33.9 | 16 |
| #40 | + | + | 25.4 | 25.2 | 2,784 | + | 15.0 | 1,073,000 |
| #41 | + | + | 23.2 | 23.6 | 8,317 | + | 24.9 | 3,272 |
| #42 | + | + | 22.7 | 23.0 | 11,610 | + | 13.7 | 3,828,000 |
| #43 | + | + | 25.7 | 25.3 | 2,482 | + | 17.0 | 1,878,000 |
| #44 | + | + | 27.2 | 27.5 | 877 | + | 33.2 | 19 |
| #45 | + | + | 27.4 | 27.4 | 865 | + | 20.7 | 32,690 |
| #46 | + | + | 17.9 | 18.0 | 184,300 | + | 14.1 | 1,510,000 |
| #47 | + | + | 26.3 | 26.4 | 1,542 | + | 13.5 | 2,127,000 |
| #48 | + | + | 17.0 | 17.1 | 310,350 | + | 16.0 | 1,195,000 |
| #49 | + | + | 23.3 | 23.3 | 8,728 | + | 13.5 | 24,140,000 |
| #50 | + | + | 24.2 | 24.5 | 4,800 | + | 20.8 | 39,270 |
| #51 | + | + | 22.3 | 22.6 | 14,150 | + | 16.3 | 1,062,000 |
| #52 | + | + | 25.1 | 25.3 | 2,993 | + | 17.7 | 441,300 |
| #53 | + | + | 27.7 | 27.5 | 747 | + | 20.3 | 156,200 |
| #54 | + | + | 20.7 | 21.1 | 35,410 | + | 21.7 | 57,670 |
| #55 | + | + | 27.3 | 27.5 | 870 | + | 18.1 | 178,000 |
| #56 | + | + | 34.4 | ND | 12 | + | 21.2 | 41,280 |
| #57 | + | + | 22.0 | 22.3 | 17,005 | + | 31.4 | 64 |
| #58 | + | + | 30.0 | 30.5 | 173 | + | 17.8 | 240,400 |
| #59 | + | + | 18.4 | 18.6 | 137,800 | + | 13.5 | 4,642,000 |
| #60 | + | + | 14.8 | 14.9 | 1,072,400 | + | 14.7 | 2,772,000 |

*Ct* cycle threshold, *ND* not detected, *NIID* National Institute of Infectious Diseases, *RT-PCR* reverse transcription polymerase chain reaction
